# Anaesthesia and infection control in cesarean section of pregnant women with coronavirus disease 2019 (COVID-19)

**DOI:** 10.1101/2020.03.23.20040394

**Authors:** LinLi Yue, Lefei Han, Qiannan Li, Min Zhong, Jun Wang, Zhenzhen Wan, Caijuan Chu, Yi Zeng, Min Peng, Lin Yang, Na Li

**Author notes:** Correspondence addressed to Dr Na Li, Department of Anesthesiology, Maternal and Child Health Hospital of Hubei Province,; and Dr Lin Yang, School of Nursing, The Hong Kong Polytechnic University, Hong Kong Special Administrative Region, China. These authors equally contributed to this study (LinLi Yue, Lefei Han, Qiannan Li).

## Abstract

**Background:** The coronavirus disease 2019 (COVID-19) first emerged in Wuhan, China, and soon caused an ongoing pandemic globally. In this study we conducted a retrospective study to evaluate the safety and efficacy of combined spinal-epidural anaesthesia (CSEA) and infection control measures on perinatal care quality of 30 pregnant women with confirmed and suspected COVID-19.

**Methods:** Individual demographic data, clinical outcomes, laboratory investigations of pregnant women and their newborns were collected from electronic medical records of the Maternal and Children Health Hospital of Hubei Province, during January 24 to February 29, 2020. Anaesthesia and surgery results were compared between pregnant women with confirmed and suspected COVID-19 infection.

**Results:** Using CSEA in cesarean section was effective and safe for pregnant women with confirmed and suspected COVID-19 infection. Administration of dezocine and morphine was effective as postoperative analgesia, and well tolerated in COVID-19 patients. The assessment of surgery outcomes also showed similar results in both confirmed and suspected cases. No respiratory failure nor distress were found in the mothers with confirmed COVID-19 infection and their neonates. None of these patients experienced severe obstetric complications related to anaesthesia and surgeries. No COVID-19 infection was reported in the neonates born to the mothers with confirmed COVID-19 infection and healthcare workers in these operations.

**Conclusions:** In cesarean section for pregnant women with COVID-19 infection, CSEA was safe and efficient in achieving satisfactory obstetrical anaesthesia and postoperative analgesia. No cross-infection occurred in the HCWs working in these operations.

## Introduction

The epidemics of 2019 coronavirus infectious disease (COVID-19) first emerged in Wuhan, China, and soon spread throughout China and more than 100 other countries within three months ^1^. As of March 4, 2020, there are cumulatively 80,424 and 12,553 cases of COVID-19 infection inside and outside China. The causative pathogen was identified as SARS-CoV-2 which was genetically related to two previous highly-pathogenic coronaviruses SARS-CoV (79% similarity) and MERS-CoV (50%) ^2^. Epidemiological studies in China have found that SARS-CoV-2 had a higher transmission rate (basic reproductive number ranges from 2 to 3)^3^, and a varying case fatality rate across age groups (range from 0 in children to 14.8% in patients aged 80 years or over) ^4^.

Although SARS-CoV and MERS-CoV have caused high mortality and severe complications in pregnant women^5,6^, current evidence did not support that SARS-CoV-2 infection caused acute respiratory distress, miscarriage, and stillbirth ^7^. The data on neonates born to the mothers with COVID-19 infection were also limited so far ^8^. However, since SARS-CoV-2 is highly contagious, it poses a big threat to healthcare workers, other pregnant women and infants.

In the COVID-19 epicenter Wuhan, a total of 49,540 cases and 2,871 deaths have been reported as of 4 March 2020. As one of the largest maternal and children hospitals in Wuhan, the Maternal and Children Health Hospital of Hubei Province has 1,500 hospital beds in total. From January 24 to March 15, 2020, there were 3,294 pregnant women who had vaginal or operative deliveries in this hospital, of whom 110 with suspected and confirmed COVID-19 infection. Given an increasing number of pregnant women in our hospital, COVID-19 infection has been as one indication for cesarean section since 24 January 2020.

Anesthesiologists had a high exposure risk to SARS-CoV-2 when conducting cesarean section for pregnant women with pneumonia. In response, we upgraded infection prevention and control practice to the highest level according to the national guideline ^9^. One negative pressure operating room (OR) was dedicated to all operations on pregnant women with confirmed COVID-19 pneumonia. The suspected cases were operated in two other operating rooms in the designated area. A series of training and monitoring procedures have been implemented in our hospital to enhance the adherence to the infection prevention and control guidelines, particularly on hand hygiene, donning and doffing of PPE, contact and airborne precautions.

To minimize the occupational risk of anesthesiologists, we made a special arrangement of using the combined spinal-epidural anaesthesia (CSEA) for COVID-19 patients, if no contraindications were indicated. According to the Practice Guidelines for Obstetric Anaesthesia by the American Society of Anesthesiologists Task Force on Obstetric Anaesthesia and the Society for Obstetric Anaesthesia and Perinatology ^10^, CSEA has the advantages of combining anaesthesia during operations with postpartum patient controlled epidural analgesia. Compared to epidural anaesthesia, CSEA can provide a more effective and rapid onset of analgesia for labor ^11^. Moreover, anaesthesia nurses are exempted from entering isolation rooms for managing epidural catheters of postpartum patients, thereby further minimizing occupation exposure risks and the chance of cross-infection.

This study aims to evaluate the safety and efficacy of CSEA and infection control measures on perinatal care quality of 30 pregnant women with confirmed and suspected COVID-19 infection.

## Methods

### Patients

Medical records of pregnant women who were admitted into our hospital for scheduled or emergency cesarean section were retrospectively retrieved during the period of January 24 – February 29, 2020. The diagnosis criteria followed the guideline by the National Health Commission of China ^12^. Suspected COVID-19 infection fulfilled at least two of following criteria: having an exposure history; having fever, lymphopenia or lower counts of WBC; typical chest CT images of COVID-19 infection (multiple ground-glass opacity). Throat swabs were collected from all suspected cases and sent to the Wuhan Provincial Centre of Disease Prevention and Control for reverse transcription polymerase chain reaction (RT-PCR) tests. COVID-19 infection was confirmed if patients were tested positive.

### Infection prevention and control

All healthcare personnel working in the negative pressure OR were required to enter the anteroom for donning the full set of personal protection equipment (PPE): N95 respirator, fluid resistant protective gown, gloves, full face shield and goggles, before entering the OR. A nursing staff stood by in anteroom to cater emergency requests and coordinate patient movement. Hypochlorite disinfectant at 2000mg/L was used for surface and floor decontamination before and after operation. Air disinfection was conducted by hydrogen peroxide air spray (1%, at 10ml/m^3^), followed by one hour disinfection of ultraviolet lights and a movable air disinfection machine. Body fluid spillover was cleaned by paper towel and then disinfected by hypochlorite disinfectant for 10-30min.

### Anaesthesia procedure

The procedure of CSEA followed a standard protocol of our hospital. After giving 500ml lactated Ringer’s solution by rapid intravenous infusion, intraspinal anaesthesia (0.5% ropivacaine at 10-15mg) was administered using pencil-point spinal needles at L2-3. An epidural catheter was inserted toward the head as a rescue pathway during operation and also for postoperative analgesia. For postoperative analgesia, dezocine (5mg/ml) at 5-10mg was immediately administered by intravenous infusion after delivery, combined with morphine (0.2mg/ml) at 2mg injected via epidural catheter at the end of caesarean section. Some patients also used patient epidural analgesia (PEA). The loading dose was morphine (0.1mg/ml) at 1mg, followed by 50ml epidural solution that contains 0.75mg/ml ropivacaine and 0.04mg/ml morphine, at a basal infusion rate of 2ml/hr.

### Data collection and analysis

The encrypted individual demographic data, clinical data, laboratory investigations, discharge or transfer records of pregnant women and their newborns were collected from electronic medical records. Descriptive statistics such as mean and standard deviation were calculated for continuous data and frequencies were calculated for categorical data. Fisher’s exact tests and Mann-Whitney U tests were conducted to compare between patients with confirmed and suspected COVID-19 infection. All analysis was conducted using the R software version 3.6.2.

## Results

### Demographic characteristics and clinical outcomes of patients

There were fourteen patients with confirmed COVID-19 infection, and sixteen suspected cases who had cesarean section during the study period. The demographic and clinical characteristics of patients are shown in Table 1. More than 70% of these patients had complications in pregnancy. Three (20.0%) confirmed cases and three (17.6%) suspected cases had pre-term delivery. No respiratory failure nor distress were found in the mothers and their neonates. No COVID-19 infection was reported in these neonates.

**Table 1.**
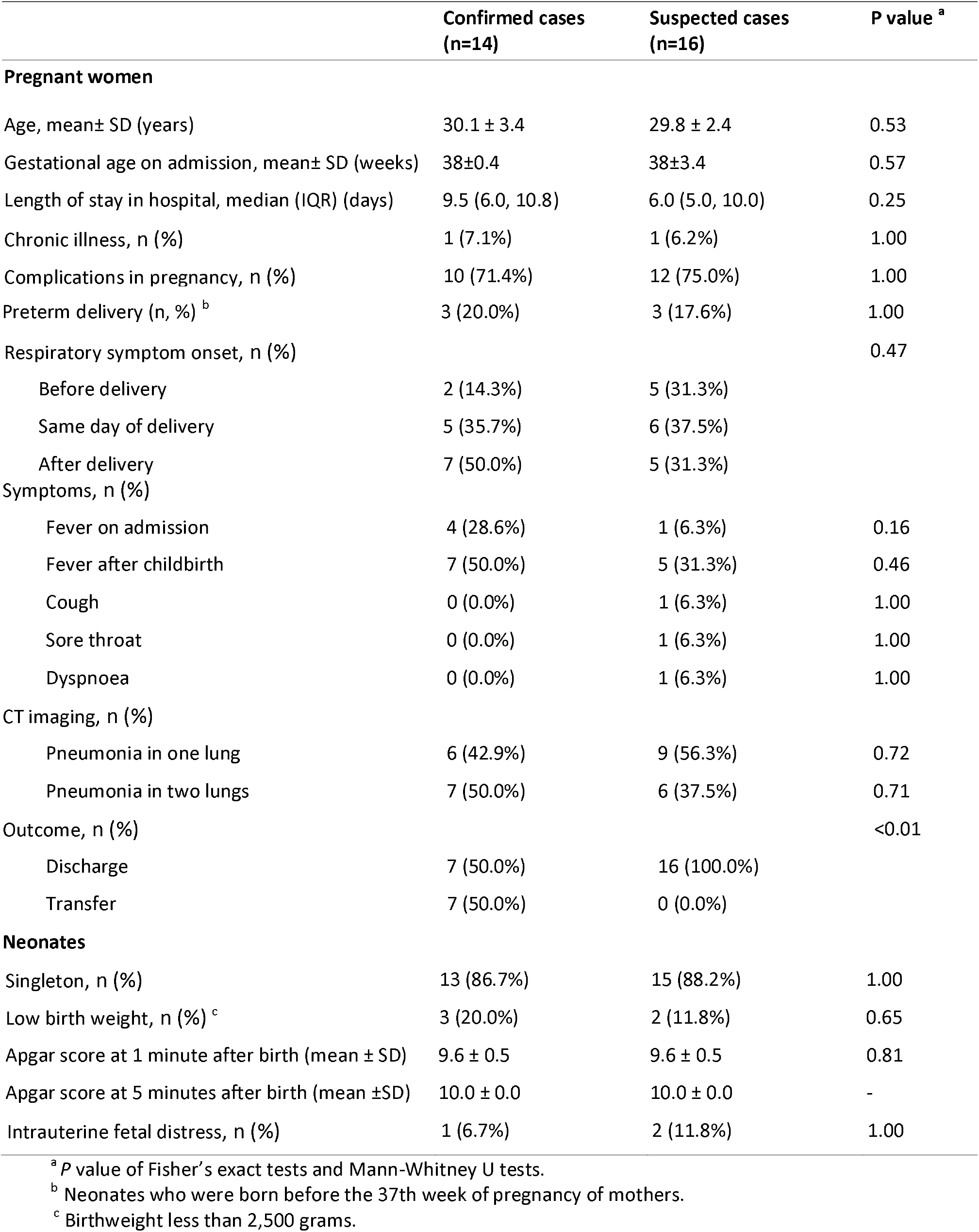
Demographic characteristics, maternal and neonatal outcomes of patients with confirmed and suspected COVID-19 infection.

|  | Confirmed cases<br>(n=14) | Suspected cases<br>(n=16) | P value <sup>a</sup> |
| --- | --- | --- | --- |
| **Pregnant women** |  |  |  |
| Age, mean± SD (years) | 30.1 ± 3.4 | 29.8 ± 2.4 | 0.53 |
| Gestational age on admission, mean± SD (weeks) | 38±0.4 | 38±3.4 | 0.57 |
| Length of stay in hospital, median (IQR) (days) | 9.5 (6.0, 10.8) | 6.0 (5.0, 10.0) | 0.25 |
| Chronic illness, n (%) | 1 (7.1%) | 1 (6.2%) | 1.00 |
| Complications in pregnancy, n (%) | 10 (71.4%) | 12 (75.0%) | 1.00 |
| Preterm delivery (n, %) <sup>b</sup> | 3 (20.0%) | 3 (17.6%) | 1.00 |
| Respiratory symptom onset, n (%) |  |  | 0.47 |
| Before delivery | 2 (14.3%) | 5 (31.3%) |  |
| Same day of delivery | 5 (35.7%) | 6 (37.5%) |  |
| After delivery | 7 (50.0%) | 5 (31.3%) |  |
| Symptoms, n (%) |  |  |  |
| Fever on admission | 4 (28.6%) | 1 (6.3%) | 0.16 |
| Fever after childbirth | 7 (50.0%) | 5 (31.3%) | 0.46 |
| Cough | 0 (0.0%) | 1 (6.3%) | 1.00 |
| Sore throat | 0 (0.0%) | 1 (6.3%) | 1.00 |
| Dyspnoea | 0 (0.0%) | 1 (6.3%) | 1.00 |
| CT imaging, n (%) |  |  |  |
| Pneumonia in one lung | 6 (42.9%) | 9 (56.3%) | 0.72 |
| Pneumonia in two lungs | 7 (50.0%) | 6 (37.5%) | 0.71 |
| Outcome, n (%) |  |  | <0.01 |
| Discharge | 7 (50.0%) | 16 (100.0%) |  |
| Transfer | 7 (50.0%) | 0 (0.0%) |  |
| **Neonates** |  |  |  |
| Singleton, n (%) | 13 (86.7%) | 15 (88.2%) | 1.00 |
| Low birth weight, n (%) <sup>c</sup> | 3 (20.0%) | 2 (11.8%) | 0.65 |
| Apgar score at 1 minute after birth (mean ± SD) | 9.6 ± 0.5 | 9.6 ± 0.5 | 0.81 |
| Apgar score at 5 minutes after birth (mean ± SD) | 10.0 ± 0.0 | 10.0 ± 0.0 | - |
| Intrauterine fetal distress, n (%) | 1 (6.7%) | 2 (11.8%) | 1.00 |

<sup>a</sup> P value of Fisher's exact tests and Mann-Whitney U tests.
<sup>b</sup> Neonates who were born before the 37th week of pregnancy of mothers.
<sup>c</sup> Birthweight less than 2,500 grams.

### Anaesthesia and surgery evaluations

Anaesthesia results are summarized in Table 2. Most patients had sensory blockade levels above T5 and Bromage scale of 3 during operation. The confirmed cases showed as good tolerance to anaesthesia as suspected ones. Four suspected COVID-19 cases, but none of confirmed cases, experienced intraoperative hypotension. None of these patients required intraoperative sedation nor airway management. The majority ranked the grade 1 in anaesthesia effect.

**Table 2.**
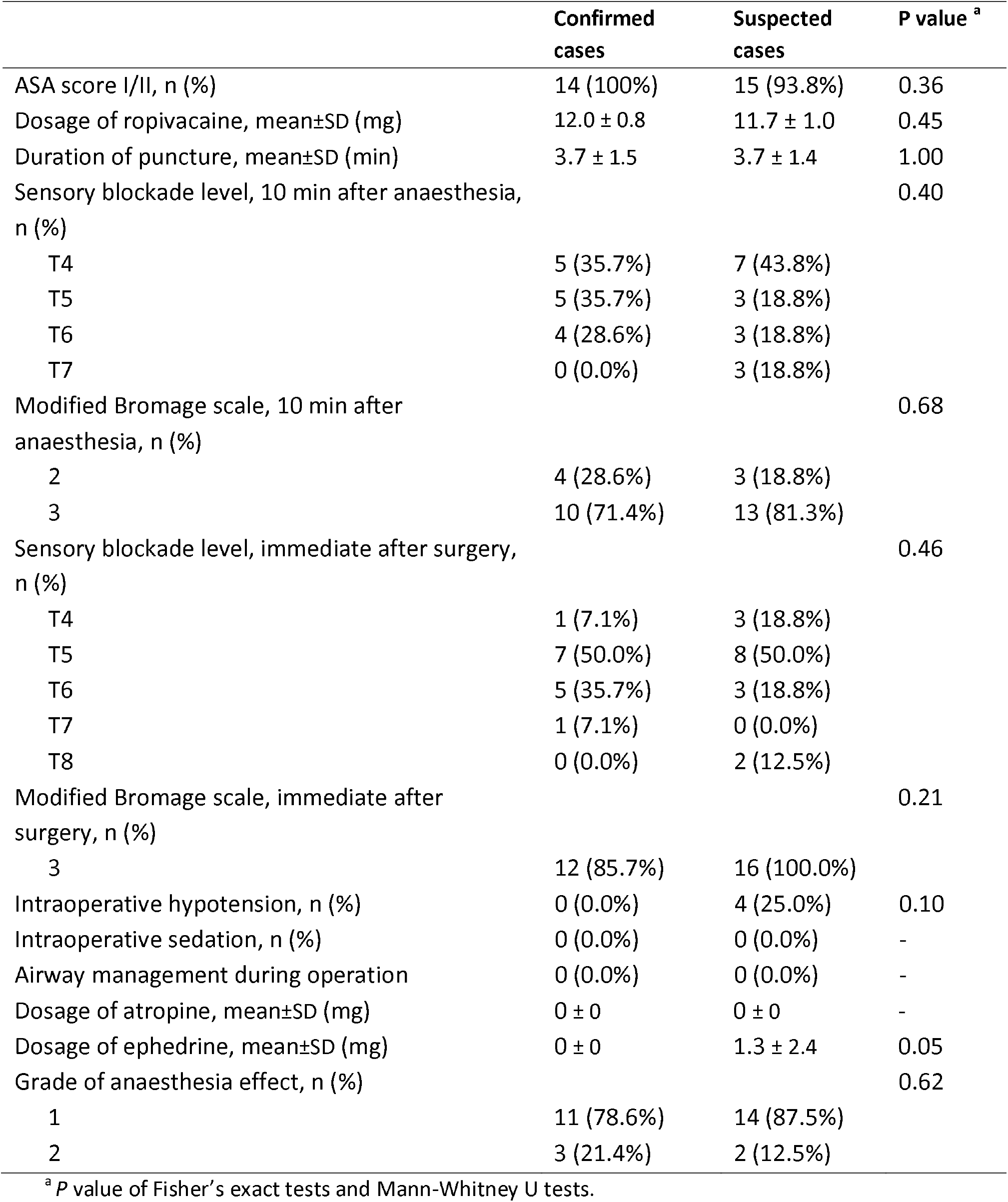
Anaesthesia evaluations in patients with confirmed and suspected COVID-19 infection.

|  | **Confirmed cases** | **Suspected cases** | **P value <sup>a</sup>** |
| --- | --- | --- | --- |
| ASA score I/II, n (%) | 14 (100%) | 15 (93.8%) | 0.36 |
| Dosage of ropivacaine, mean±SD (mg) | 12.0 ± 0.8 | 11.7 ± 1.0 | 0.45 |
| Duration of puncture, mean±SD (min) | 3.7 ± 1.5 | 3.7 ± 1.4 | 1.00 |
| Sensory blockade level, 10 min after anaesthesia, n (%) |  |  | 0.40 |
| T4 | 5 (35.7%) | 7 (43.8%) |  |
| T5 | 5 (35.7%) | 3 (18.8%) |  |
| T6 | 4 (28.6%) | 3 (18.8%) |  |
| T7 | 0 (0.0%) | 3 (18.8%) |  |
| Modified Bromage scale, 10 min after anaesthesia, n (%) |  |  | 0.68 |
| 2 | 4 (28.6%) | 3 (18.8%) |  |
| 3 | 10 (71.4%) | 13 (81.3%) |  |
| Sensory blockade level, immediate after surgery, n (%) |  |  | 0.46 |
| T4 | 1 (7.1%) | 3 (18.8%) |  |
| T5 | 7 (50.0%) | 8 (50.0%) |  |
| T6 | 5 (35.7%) | 3 (18.8%) |  |
| T7 | 1 (7.1%) | 0 (0.0%) |  |
| T8 | 0 (0.0%) | 2 (12.5%) |  |
| Modified Bromage scale, immediate after surgery, n (%) |  |  | 0.21 |
| 3 | 12 (85.7%) | 16 (100.0%) |  |
| Intraoperative hypotension, n (%) | 0 (0.0%) | 4 (25.0%) | 0.10 |
| Intraoperative sedation, n (%) | 0 (0.0%) | 0 (0.0%) | - |
| Airway management during operation | 0 (0.0%) | 0 (0.0%) | - |
| Dosage of atropine, mean±SD (mg) | 0 ± 0 | 0 ± 0 | - |
| Dosage of ephedrine, mean±SD (mg) | 0 ± 0 | 1.3 ± 2.4 | 0.05 |
| Grade of anaesthesia effect, n (%) |  |  | 0.62 |
| 1 | 11 (78.6%) | 14 (87.5%) |  |
| 2 | 3 (21.4%) | 2 (12.5%) |  |

<sup>a</sup> P value of Fisher's exact tests and Mann-Whitney U tests.

The mean duration of surgery was 39.4 and 37.3 minutes for confirmed and suspected cases, respectively (Table 3). It took an average of 3.7 minutes for anesthetists to complete puncture. The patients with confirmed COVID-19 infection had more bleeding and longer time to anal exhaust. The other results were similar between two groups.

**Table 3.** Surgery evaluations in patients with confirmed and suspected COVID-19 infection.

|  | Confirmed cases | Suspected cases | P value <sup>a</sup> |
| --- | --- | --- | --- |
| Duration of surgery, mean±SD (min) | 39.4 ± 8.9 | 37.3 ± 5.2 | 0.53 |
| Volume of bleeding, mean±SD (ml) | 335.7 ± 63.3 | 293.8 ± 25.0 | 0.02 |
| Volume of urine, mean±SD (ml) | 307.1 ± 64.6 | 281.3 ± 155.9 | 0.08 |
| Volume of fluid infusion, mean±SD (ml) | 1342.0 ± 295.4 | 1281.3 ± 268.9 | 0.81 |
| Anal exhaust time, mean±SD (hour) | 20.7 ± 4.2 | 16.5 ± 4.4 | 0.03 |
| Time to straight leg raising, mean±SD (hour) | 3.3 ± 1.3 | 2.9 ± 1.1 | 0.56 |
| Dezocine (IV)+ Morphine (epidural), n (%) | 6 (42.9%) | 5 (31.3%) | 0.71 |
| Patient epidural analgesia, n (%) | 7 (87.5%) | 4 (80.0%) | 1.00 |

<sup>a</sup> P value of Fisher's exact tests and Mann-Whitney U tests.

### Safety and side effects

The vital signs of patients remained stable during the surgery (Figure 1). Both groups achieved satisfactory visual analogue scale (VAS) and Bruggrmann Comfort Scale scores (BCS). Some minor complications were reported in these patients, including itchiness (n=1, probably due to allergy to Morphine), postoperative vomit and nausea (n=2, probably caused by analgesia medicine), pain at anaesthesia injection site (n=3). None of the patients reported the incidence of severe obstetric complications related to anaesthesia and surgeries, such as incision wound infection, postpartum haemorrhage, intestinal adhesion, intestinal obstruction, or venous thrombosis. No events of atelectasis or hypoxia occurred in these patients.

**Figure 1.**
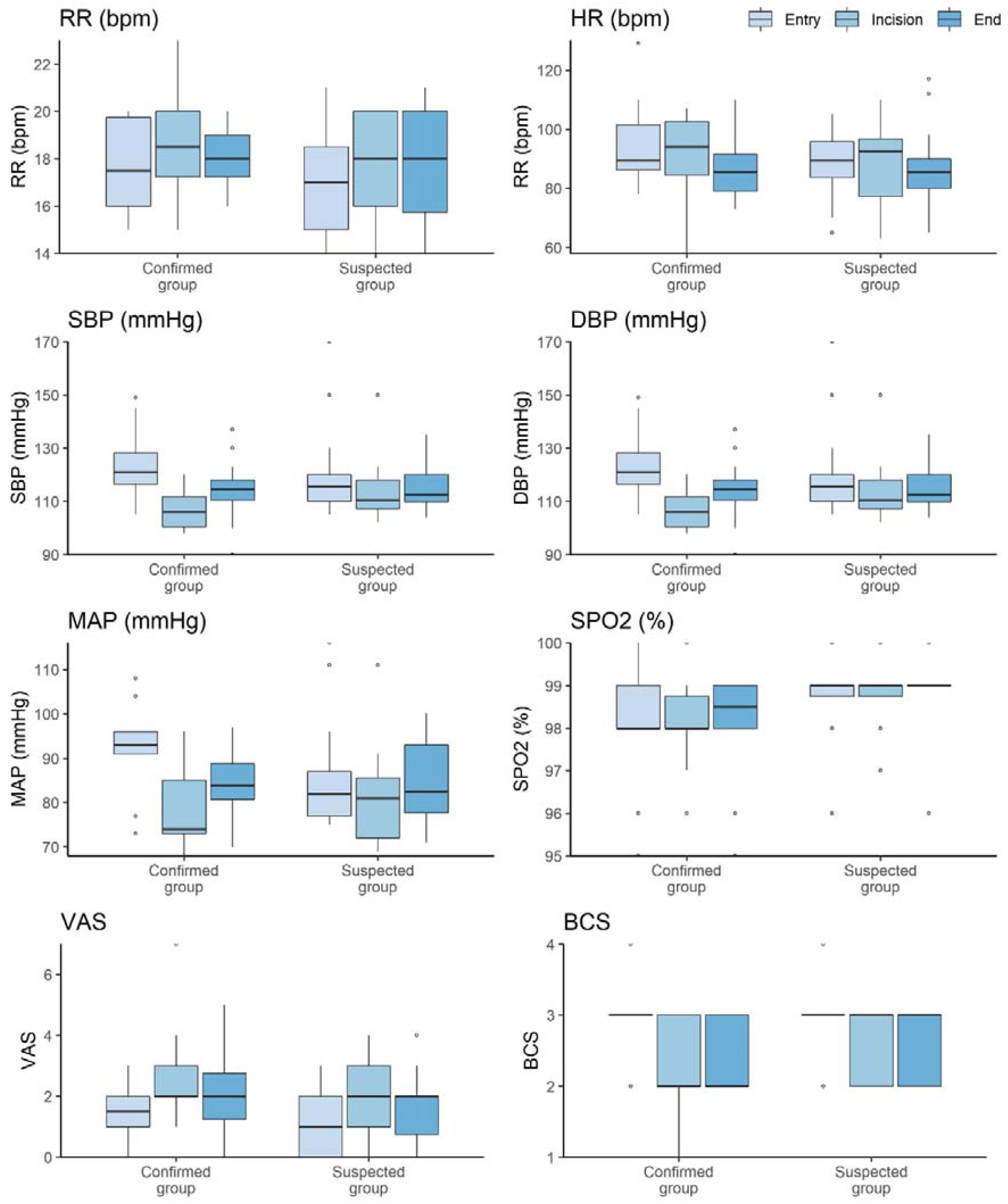
Anaesthesia assessments during cesarean section, at time of patient entry (light blue), incision (blue) and the end of operation (dark blue). Abbreviations: RR, respiratory rate; HR, heart rate; SBP, systolic blood pressure; DBP, dialytic blood pressure; MAP, mean arterial pressure; SPO2, saturation percentage of oxygen; VAS, visual analogue scale; BCS, Bruggrmann Comfort Scale scores.

## Discussion

In this retrospective study, we found that the CSEA generally achieved a satisfactory performance and was a safe alternative to epidural anaesthesia we usually used in non-contagious mothers. As of 13 March 2020, none of our staff at the Anaesthesia Department were diagnosed with COVID-19 infection. No vertical transmission of COVID-19 was found in the neonates born to the mothers with confirmed infection, which was consistent with a recent case report in another tertiary hospital of Wuhan ^7^.

Favre et al suggested that when possible, vaginal delivery should be preferred over surgical delivery in pregnant women with COVID-19 infection to reduce the risk of severe maternal complications ^13^. Our study showed that cesarean section with CSEA was safe for contagious pre-laboring women with suspected or confirmed COVID-19 infection, in either scheduled or emergency surgeries. We found that COVID-19 infection caused relatively mild symptoms in pregnant women, who rarely had hypoxia or respiratory failure. The maternal and neonatal outcomes were also comparable to other pregnant women without infection ^14^. Compared to cesarean section, the exposure risk of HCWs inside labor room could increase to a greater extent due to longer duration of vaginal delivery. The lessons from the previous SARS outbreak also found that the GA was a high-risk procedure for both HCWs and patients. One previous study in Canada reported that one critical care doctor infected SARS-CoV when conducting a prolonged intubation procedure for GA of SARS patients, despite that this doctor was wearing an N95 respirator and goggles ^15^.

Given the high transmissibility of SARS-CoV-2 ^3, 16^, contact and airborne precautions have been stringently enforced in the operation theaters designated to patients with confirmed or suspected COVID-19 infection in our hospital. However, the highest level of infection control measures could have prompted the adverse emotional response of pre-laboring mothers. Anxiety and stress have been widely reported in Wuhan residents ^17^. Pregnant women might also worry about health status of their newborns, although there is limited evidence to suggest a vertical transmission could occur in COVID patients ^7, 14, 18^. The CSEA could have provided rapid onset of analgesia during cesarean section, which could benefit patients by effectively shorten the surgery period and partially relieve mental stress of patients.

Early clinical reports of COVID-19 infection in general populations have found fever and cough was the most common symptoms on admission ^19^. By contrast, less than 30% of pregnant women with confirmed COVID-19 presented fever on admission, and about 20% did not develop fever during hospital stay. This highlights the difficulty of early detection of the potentially contagious patients in obstetric wards. Interestingly, over 85% of them (12 out of 14) already had abnormal chest CT imaging in single lungs or both lungs on admission. Hence, in the absence of point-of-care rapid reverse transcription polymerase chain reaction (RT-PCR) tests, low-dose chest CT scan could be used as an alternative screening tool for COVID-19 infection.

The infection prevention and control guidelines in our hospital are in line with those recently proposed by Peng *et al*. based on the previous experience of the 2003 SARS outbreak ^19^. Although SARS-CoV-2 had a lower case fatality ratio than SARS-CoV and MERS-CoV, its transmissibility is much higher than previous coronaviruses ^16^. As of February 11,2020, there were 1716 HCWs infected by SARS-CoV-2, of whom 14.8% were severe or critical cases and five deaths reported ^4^. Therefore, strict adherence to the infection prevention and control guidelines is critical to reduce the risk of cross infections between HCWs and patients. Our hospital has held regular online trainings and practical workshops to all HCWs and supporting staff. Wearing the full set of PPE did not affect the performance of anaesthesia and effectively protected the frontline staff from occupational exposure to COVID-19 infections. It is of note that even in countries with relatively small numbers of cases, HCWs still need to maintain high vigilance against this virus, given the RT-PCR test capacity might be limited in some areas and cases are seriously underreported at the early stage of the epidemic ^20^. As the epidemic progresses, an exponential increase of patient numbers could cause a serious shortage of PPE in healthcare settings.

Our study has a few limitations. First, we have a small sample size of 34 pregnant women, although this is the largest sample for pregnant women with COVID-19 infection to date. Second, all patients were in the third trimester. All of them had relatively mild infection and most did not show any respiratory symptoms on admission. Hence, it is not clear that our findings can also be applied to patients with moderate and severe infection, since they might require endotracheal intubation or other airway management during surgery. Last but not least, this is a one-center retrospective study at the epicenter of COVID outbreaks, so the results might be generalized to other settings.

## Conclusions

In cesarean section for pregnant women with COVID-19 infection, CSEA was safe and efficient in achieving satisfactory obstetrical anaesthesia and could also assist administration of dezocine and morphine for postoperative analgesia. No adverse events associated with anaesthesia and surgery were found in these patients, and no cross-infection occurred in the HCWs working in these operations.

## Authors’ Contributions

NL and LYang initiated and designed the study. MZ, JW, ZW, CC and YZ contributed to data collection and clean. LH conducted data analysis. LYue, LH, QL, NL and LYang interpreted the findings and drafted the manuscript. All the authors proved the final version of this manuscript.

## Data Availability

All data and materials used in this work were available based on request.

## Ethical consideration

The ethical approval has been obtained from the Ethics Committee of the Maternal and Child Health Hospital of Hubei Province [2020-IEC-LW011].

